# High serum uric acid levels are associated with worse cognitive performance: A cross-sectional study

**DOI:** 10.1101/2022.09.11.22279827

**Authors:** Yousef Khaled, Aya A. Abdelhamid, Hissa Al-Mazroey, Abdulrahman K. Almannai, Sara Fetais, Aisha S. Al-Srami, Shaima Ahmed, Noora Al-Hajri, Tawanda Chivese, Laiche Djouhri

## Abstract

**Background:** The association between raised serum uric acid and cognitive dysfunction remains a debated issue. In the present study, we investigated whether serum uric acid is associated with cognitive performance in a cohort of healthy individuals in Qatar;

**Methods:** We conducted a cross-sectional study on a cohort of individuals aged 40 to 80 years old, without a diagnosis of dementia, schizophrenia, and stroke, who participated in Qatar Biobank. Cognitive performance was assessed using the CANTAB’s paired episodic memory test and reaction time test. The participants were divided into two groups, one group with a normal serum uric acid level (<350 μmol/l) and the other with high serum uric acid levels (>350 μmol/l). Two multivariable linear regression models were applied to determine the association between serum uric acid and memory test performance score and between serum uric acid and reaction time test performance score;

**Results:** A total of 996 individuals with median age 48.0 years, IQR: (44.5, 54.0), of which 48.2% were male, were included. After adjusted multivariable linear regression, high serum uric acid levels were strongly associated with poor performance in the paired episodic memory test (beta -6.25, 95%CI - 10.65 to -1.84, p= 0.006). However, after adjusted multivariable linear regression, we found no significant association between high serum uric acid and performance in the reaction time test (beta - 13.24, 95%CI -138.77 to 112.29, p= 0.836,);

**Conclusions:** In a cohort of healthy individuals in Qatar, high serum uric acid levels are associated with worse performance in visual memory and new learning domains of cognitive function but no significant effect on processing speed function.

## 1. Introduction

Uric acid (UA) is a natural waste product from the digestion of purine-rich foods that are generally abundant in high-protein diets [1]. UA plays an essential role in human metabolism [2] and is a major natural antioxidant that prohibits the occurrence of cellular damage and accounts for a considerable part of the antioxidative capacity of the plasma [3]. Several studies have investigated the physio-pathological role of UA in cognitive function by measuring serum UA (sUA) levels which represent the balance between dietary purine intake, xanthine oxidase activity, and renal UA excretion [4]. Although those studies examined the association between sUA with various cognitive outcomes among both middle-aged and older adults [5-15], most of them focused on the elderly population. Some of those studies suggest that UA has a beneficial effect on cognitive performance or a protective role against the progression of cognitive impairment [12-16], whereas other studies reached the opposite conclusion and reported a potentially adverse effect of hyperuricemia on cognitive outcomes and that UA was a risk factor for cognitive function [6-12,17]. Thus, the relationship between UA and cognitive function remains a debated issue that warrants further investigations.

Conflicting results have also been reported in studies investigating the effects of UA on cognitive function in patients with different types of dementia. For example, Latourte and co-workers [18] suggested that high sUA levels constitute a potential risk factor for dementia. In contrast, the findings of an earlier study [13] suggested that UA might reduce the risk of dementia and cognitive impairment. sUA levels were also found to be lower in patients with Alzheimer’s disease and in subjects with mild cognitive impairment than those in healthy controls, suggesting that UA may have a protective effect [19]. Thus, high levels of plasma UA might reduce oxidative stress and protect against cognitive impairment. As far as we know, there has been only one small study [7] that has analyzed the relationship between sUA and cognitive functioning in healthy elderly American adults. These investigators reported that participants with mildly elevated sUA were more likely to score in the lowest quartile of the sample on measures of cognitive functions including processing speed, verbal memory, and working memory. However, there have been no studies that have investigated the relation between sUA levels and cognitive functions in healthy participants in Qatar. Therefore, the aim of this study was to examine whether there is an association between sUA levels and cognitive performance in a cohort of healthy participants aged 40-80 years old.

## 2. Materials and Methods

### 2.1. Study design

We conducted a cross sectional study using data from the Qatar Biobank, collected during the period of 2014 to 2020. The Qatar biobank recruits citizens and long-term residents in Qatar to participate in a longitudinal observational cohort [20]. From this cohort, a sample who had uric acid measurement and cognitive assessment were randomly selected into the present study.

### 2.2. Study population

Participants included in the present study were adults aged 40 to 80 years old from the QBB. We excluded participants with history of stroke and dementia, and who did not have either serum UA levels measured or did not perform the cognitive test. Given that stroke can cause development of dementia or cognitive impairment [21] and is associated with high serum UA levels [19], any participants with history of stroke were also excluded.

### 2.3. Data

The data were provided by the Qatar biobank (QBB) which is a national resource of collected samples and information on different aspects of health and lifestyle of many volunteers from the population of Qatar. The data included demographics, laboratory data including serum uric acid, cognitive function tests, and personal medical history.

### 2.4. Cognitive function tests

Two tests were used to assess cognitive function:

#### 2.4.1. Assessment of visual memory and new learning-paired episodic memory test Each participant carried out a memory test designed by the Cambridge Neuropsy

chological Test Automated Battery (CANTAB) [22]. The paired episodic memory test assesses visual memory and new learning. The test involves the participants learning the position of an increasing number of cards. The test uses a paired associate learning task in which a series of cards with different patterns are opened randomly and had to be learned within a short time period. The cards are then closed, and patterns are displayed one at a time in the middle of the screen. The participant must select the card that originally contains the pattern. If the participant chose the wrong card, the position of each card is presented consecutively, but not sequentially, each for two seconds. The location of each has to be recalled. The number of attempts needed to correctly identify the location of each card is summed to achieve a total guess. The time the participant spends from when the pattern is presented in the middle of screen until he selects a card is recorded as response duration. The test had a total of seven levels with the first level having two cards and the seventh level having eight cards. Participants proceeded to the next level only after successfully completing the previous level.

The outcomes for each participant were the number of levels reached, the total guesses and the time taken.

#### 2.4.2. Assessment of processing speed- reaction time test

This test, also from the CANTAB battery, assesses processing speed using a two-choice task. The test comprises of 60 trials per participant. The task generates 60 presentations of one of two targets. The target is presented as a small white box within one of two larger black boxes. The location of the target within the black box varies. During each trial, the participant has to select the box where the target appears as quickly as possible. The outcomes were the total time taken during all 60 trials, and the number of mistakes made while performing the test.

### 2.5. Other data collected

Data on gender, age, nationality, years of education, and smoking of the participants were obtained through a self-reported questioner that included detailed questions about socio-demographic factors, health conditions, and smoking habits. For participants who provided samples of blood, serum UA levels with other clinical biomarkers like free thyroxine, c-reactive protein, homocysteine, and vitamin D were measured in the laboratories of Hamad Medical Centre Laboratory in Doha. Uric acid was measured using the Enzymatic colorimetric test (Uricase). BMI was calculated after collecting various physical and clinical measurements from each participant.

### 2.6. Data Analysis

Participants were divided according to their serum UA levels into high and normal UA groups following a cut-point of 350 μmol/l [7]. The participants’ characteristics were described for the entire sample and for the two groups defined according to serum UA levels. The descriptive data were first explored for normality using histogram and the Shapiro-Wilk test. Continuous variables were described as means ± standard deviations (SD) or as median and quartiles if the assumptions of normality were not met.

Statistical tests were done to compare individuals with normal and high UA. The categorical variables were compared using the Chi-squared test. Numerical data was compared either using the t-test or the Mann–Whitney U test, if not normally distributed.

For the paired episodic memory test, the best performance in this test was assessed by completing all seven levels with least sum of total guesses and the least time taken for all levels, and vice versa the worst performance. Participants were ranked from best performance to the worst performance. The memory performance score was assigned in such a way that the highest performer had a score of 100 and the lowest performer had a score of zero.

For the reaction time test, we generated a score by the product of the sum of response duration and the sum of mistakes made across the 60 trials. The higher scores imply the worst performance and vice versa.

We compared cognitive function across the high and normal UA groups using two multivariable linear regressions, one for each test. In the first regression, the performance variable of the paired episodic memory test was the dependent variable and categorized UA was the independent variable. In the second regression, the score (product of sum of response and sum of mistakes) of the reaction time test was the dependent variable and categorized UA was the independent variable. Based on evidence from previous studies we adjusted both regressions for age [23,24], BMI [25,26], gender, and education [27,28].

All analyses were performed using the STATA statistical package, version 16.0 (StataCorp, College Station, Texas, USA).

## 3. Results

### 3.1. Demographic and clinical characteristics

We included 996 participants, 223 with high UA and 773 with normal UA. Their clinical and demographic characteristics are shown in Table 1. This sample were mostly Qatari (n= 877, 88.1%). The median age was 48.0 years (IQR 44.5, 54.0) with no differences between individuals with high and those with normal UA. Most (85.2%) of those with high uric acid were males (Table1).

**Table 1:**
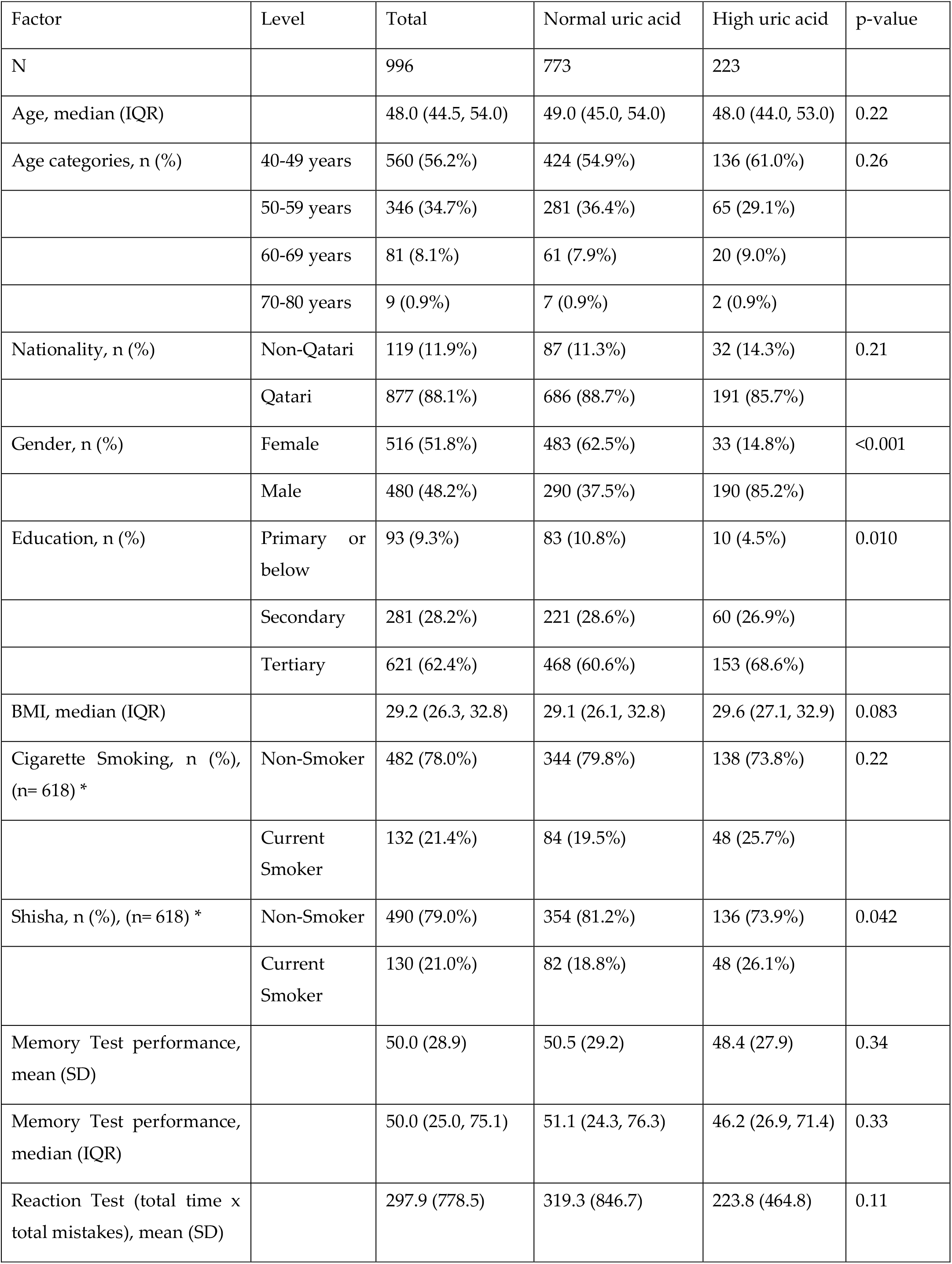
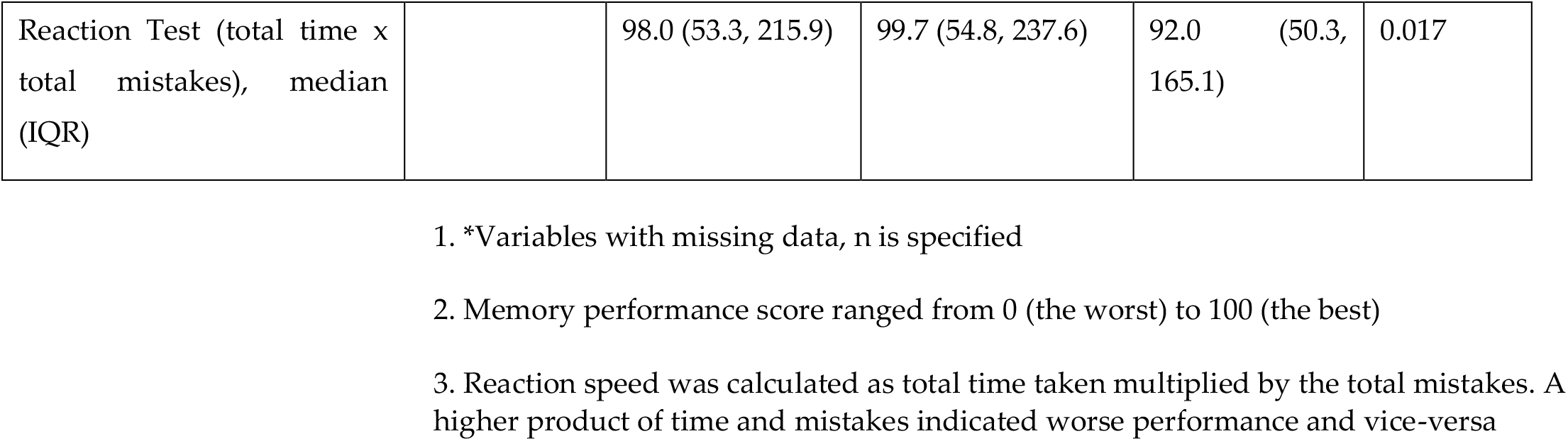
Demographic and clinical characteristics of participants

| Factor | Level | Total | Normal uric acid | High uric acid | p-value |
| --- | --- | --- | --- | --- | --- |
| N |  | 996 | 773 | 223 |  |
| Age, median (IQR) |  | 48.0 (44.5, 54.0) | 49.0 (45.0, 54.0) | 48.0 (44.0, 53.0) | 0.22 |
| Age categories, n (%) | 40-49 years | 560 (56.2%) | 424 (54.9%) | 136 (61.0%) | 0.26 |
|  | 50-59 years | 346 (34.7%) | 281 (36.4%) | 65 (29.1%) |  |
|  | 60-69 years | 81 (8.1%) | 61 (7.9%) | 20 (9.0%) |  |
|  | 70-80 years | 9 (0.9%) | 7 (0.9%) | 2 (0.9%) |  |
| Nationality, n (%) | Non-Qatari | 119 (11.9%) | 87 (11.3%) | 32 (14.3%) | 0.21 |
|  | Qatari | 877 (88.1%) | 686 (88.7%) | 191 (85.7%) |  |
| Gender, n (%) | Female | 516 (51.8%) | 483 (62.5%) | 33 (14.8%) | <0.001 |
|  | Male | 480 (48.2%) | 290 (37.5%) | 190 (85.2%) |  |
| Education, n (%) | Primary or below | 93 (9.3%) | 83 (10.8%) | 10 (4.5%) | 0.010 |
|  | Secondary | 281 (28.2%) | 221 (28.6%) | 60 (26.9%) |  |
|  | Tertiary | 621 (62.4%) | 468 (60.6%) | 153 (68.6%) |  |
| BMI, median (IQR) |  | 29.2 (26.3, 32.8) | 29.1 (26.1, 32.8) | 29.6 (27.1, 32.9) | 0.083 |
| Cigarette Smoking, n (%), (n= 618) * | Non-Smoker | 482 (78.0%) | 344 (79.8%) | 138 (73.8%) | 0.22 |
|  | Current Smoker | 132 (21.4%) | 84 (19.5%) | 48 (25.7%) |  |
| Shisha, n (%), (n= 618) * | Non-Smoker | 490 (79.0%) | 354 (81.2%) | 136 (73.9%) | 0.042 |
|  | Current Smoker | 130 (21.0%) | 82 (18.8%) | 48 (26.1%) |  |
| Memory Test performance, mean (SD) |  | 50.0 (28.9) | 50.5 (29.2) | 48.4 (27.9) | 0.34 |
| Memory Test performance, median (IQR) |  | 50.0 (25.0, 75.1) | 51.1 (24.3, 76.3) | 46.2 (26.9, 71.4) | 0.33 |
| Reaction Test (total time x total mistakes), mean (SD) |  | 297.9 (778.5) | 319.3 (846.7) | 223.8 (464.8) | 0.11 |
| Reaction Test (total time x total mistakes), median (IQR) |  | 98.0 (53.3, 215.9) | 99.7 (54.8, 237.6) | 92.0 (50.3, 165.1) | 0.017 |

1. \*Variables with missing data, n is specified

### 3.2. Comparison of cognitive function by uric acid level

In the paired episodic memory test, participants with high UA had a worse mean performance score in memory test than those with normal UA, although with weak evidence against the null hypothesis (48.4 (SD= 27.9) vs 50.5 (SD= 29.2), p=0.34). In the reaction time test, participants with high UA had lesser mean score compared to normal UA group, i.e., better performance, again with weak evidence against the null hypothesis, (223.8 (SD= 464.8) vs. 319.3 (SD= 846.7), respectively, p = 0.11) (Table1). Figure 1 shows the comparison of paired episodic test performance and reaction time test performance across both groups.

**Figure 1:**
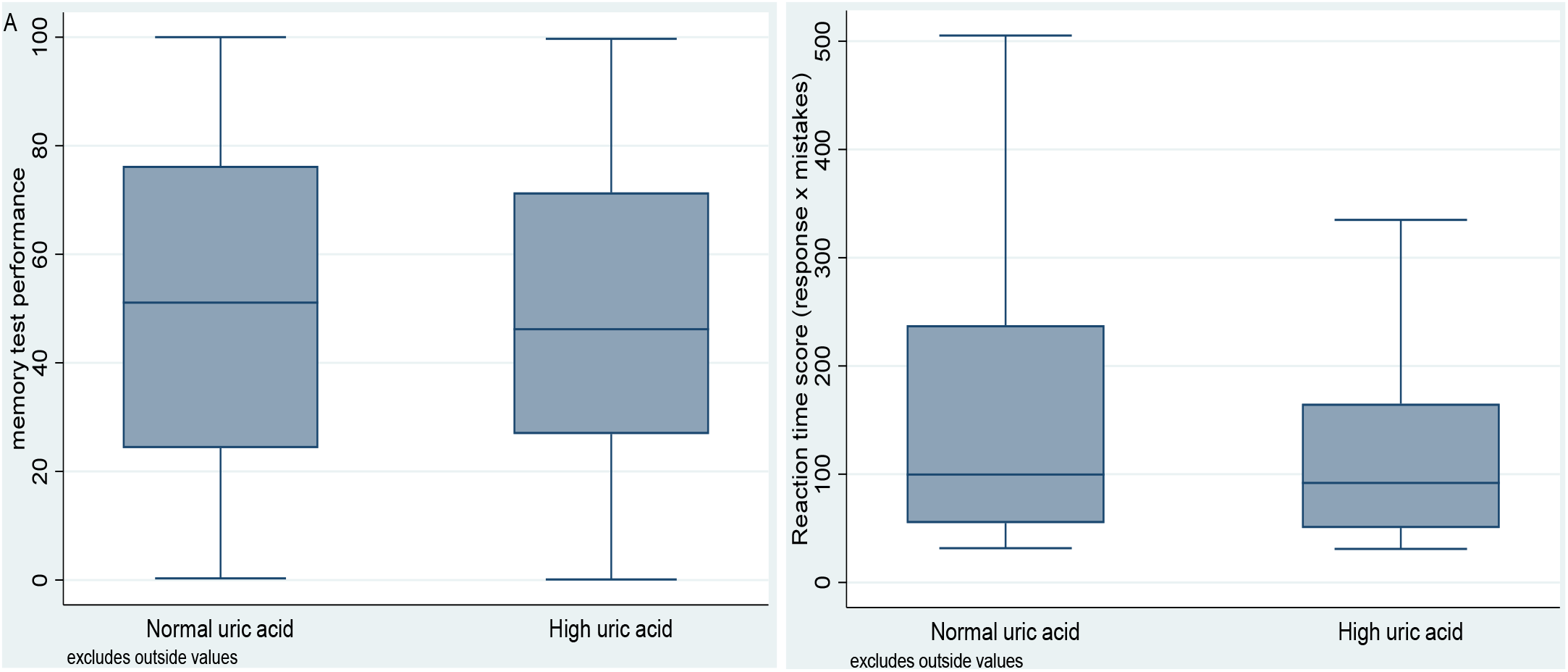
box plots comparing performance in memory test (A) and performance in reaction test (B) across normal and high uric acid groups. 1. Memory performance score ranged from 0 (the worst) to 100 (the best) 2. Reaction speed was calculated as total time taken multiplied by the total mistakes. A higher product of time and mistakes indicated worse performance and vice-versa

### 3.3. Association between high uric acid level and episodic memory and reaction time - multivariable linear regression

After multivariable linear regression, high uric acid was associated with a decrease of 6.25 in the mean performance score in the paired episodic memory test (beta -6.25, 95%CI -10.65 to -1.84, p= 0.006). However, there was no significant association between reaction time and high levels of UA (beta 13.24, 95%CI -138.77 to 112.29, p= 0.836). These findings indicate that high uric acid has significant adverse effects on the visual memory and new learning domains of cognitive function but no significant effect on processing speed function.

**Table 2:** Multivariable linear regression for association between high uric acid level and episodic memory and reaction time

|  | Coefficient | p-value | 95% Confidence interval |
| --- | --- | --- | --- |
| Memory test: performance |  |  |  |
| High UA | -6.247088 | 0.006 | -10.65396 to -1.840217 |
| Constant | 46.37756 | 0.000 | 33.97717 to 58.77796 |
| Reaction test: score |  |  |  |
| High UA | -13.23973 | 0.836 | -138.7691 to 112.2896 |
| Constant | 544.1344 | 0.003 | 190.9101 to 897.3587 |

The regression was adjusted for age, BMI, gender, and education

## 4. Discussion

In a cohort of healthy middle and old aged adults in Qatar, without existing neurocognitive diseases, asymptomatic high levels of uric acid were associated with poor visual memory and new learning function. However, high levels of uric acid were not associated with processing speed.

In this study, participants with asymptomatic high uric acid levels scored a mean of six points lower than that of the performance in visual memory and new learning of those with normal uric acid levels, after adjusting for important confounders. However, although participants of higher levels of uric acid showed better processing speed, the association went away after adjusting for confounders. The association between sUA and cognition remains controversial. Our findings were in agreement with those of other studies which concluded that UA was a risk factor for cognitive dysfunction [7,18]. Schretlen and co-workers [7] reported that participants with mildly elevated serum UA were more likely to score in the lowest quartile of the sample on measurements of cognitive functions including processing speed, verbal memory, and working memory. In addition, Latourte and coworkers after following a cohort of 1598 healthy older people for 12 years, they found an association between high sUA and increased risk of dementia. The association was more profound with vascular and mixed dementia compared to Alzheimer’s disease. However, other researchers have reported conflicting findings [13] who found that UA did not cause cognitive impairment, but instead it played a protective role. Indeed, they found that after 11 years of follow-up, high UA levels at baseline were associated with better cognitive function in healthy individuals. Low sUA levels were also reported in patients with Alzheimer’s disease [19].

The apparent discrepancy between these studies may be related to the differences between the types of cognitive tests used and/or other factors such as the age and characteristics of the participants. Furthermore, the differences between studies might be related to the fact that UA does not only have antioxidant properties, but also can acquire prooxidant attributes in some cases that might lead to damage in the vascular endothelium [29] increasing the odds of developing vascular dementia leading to worsened cognitive abilities.

The precise mechanisms by which UA could influence cognitive function remain unknown. However, several factors are believed to be involved in the pathogenesis of cognitive impairment including oxidative stress that might result from chronic exposure to environmental toxins, low antioxidant levels in the brain free radical scavenging enzymes, or mitochondrial dysfunction [4]. sUA levels were found to be associated with UA from cerebrospinal fluid, which suggests an influence of UA on brain and cognitive system [30]. It was suggested that UA which is capable of increasing the plasma antioxidant capacity (28) can promote defensive mechanisms against lipid peroxidation in the brain and promote some neuroprotective effects. However, UA also harbors prooxidant effects as it stimulates hydrogen peroxide production. In addition, catabolism of xanthine by which UA is generated produces superoxide anions. These two effects can increase the oxidative stress which will result in cell damage and apoptosis [31]. In addition, Tian and colleagues found that sUA has significant negative correlation with superoxide dismutase level, indicating the effect of UA in increasing the oxidative stress. They also found that β-Amyloid peptide level, the main component of the amyloid plaque, a well-established factor of Alzheimer’s disease pathogenesis, was positively correlated with the sUA level [31] further supporting the harmful effect of UA on cognition.

The strengths of our study include its population-based design. The sample was randomly selected from QBB, this reduced the risk of selection bias. Further, cognitive function was assessed using CANTAB tool. Because it is a nonverbal battery, it is independent of cultural differences and was found superior to language tests in detecting cognitive decline [32]. In addition, we used the paired episodic memory test, which is proven to be a sensitive test for cognitive decline [33,34].

Our study had some limitations. Due to the cross-sectional nature of the study design, temporality cannot be affirmed from this study. We could only investigate the association between UA and two domains of CANTAB, the speed of reaction and the paired memory test, but not the other cognitive function domains due to the unavailability of data. Future research should investigate the association between high UA and more domains of cognitive function, preferably in longitudinal cohorts.

## 5. Conclusions

In a cohort of healthy individuals in Qatar, high serum uric acid levels are associated with worse performance in visual memory and new learning domains of cognitive function but no significant effect on processing speed function. Further studies are required to clarify the role of uric acid in the pathogenesis of different types of dementia, and its clinical-prognostic significance.

## Data Availability

All data produced in the present work are contained in the manuscript

## Author Contributions

Conceptualization, Y.K.; methodology, Y.K., A A.A., H.A., T.C. and L.D.; formal analysis, Y.K., A A.A. and T.C.; data curation, Y.K., A A.A. and T.C.; writing—original draft preparation, Y.K., A A.A., H.A., A K.A., S.F., A S.A., S.A. and N.A.; writing—review and editing, Y.K., A A.A., T.C. and L.D.; visualization, A A.A.; supervision, T.C. and L.D. All authors have read and agreed to the published version of the manuscript.

## Institutional Review Board Statement

Ethical approval was obtained from the institutional review board of Qatar Biobank (Ex -2019-RES-ACC-0182-0107) as well as Qatar University institutional review board (QU-IRB) (QU-IRB 1223 E/20).

## Informed Consent Statement

Informed consent was obtained from all subjects involved in the study.

## Data Availability Statement

The data in this study was obtained from Qatar Biobank where restrictions may apply. Such dataset may be requested from.

## Acknowledgments

We thank Qatar Biobank for providing the data. We also thank the Population Medicine Department at Qatar University for methodological and statistical support.

